# Determinants of Sense of Coherence among older adults attending a Geriatric Centre in Nigeria A study Protocol

**DOI:** 10.1101/2023.06.20.23291667

**Authors:** Lawrence Adekunle Adebusoye, Oluwagbemiga Oyinlola, Eniola Olubukola Cadmus

## Abstract

**Background:** Old age is a stage of life in which people face changes in their physical and psycho-emotional aspects. Thus, having an adequate sense of coherence (SOC) is required to face these situations successfully. The SOC (comprehensibility, manageability, and meaningfulness of life) is defined as a global orientation expressing a person’s pervasive and enduring feeling of confidence modified by stimuli derived from one’s internal and external environments while living, the resources available to meet the demands posed by these stimuli, and the fact that these demands are challenges worthy of investment and engagement. Empirical evidence on the SOC available to older persons is lacking in countries like Nigeria. This study aims to investigate the Sense of Coherence (SOC) available to older patients attending the Chief Tony Anenih Geriatric Centre (CTAGC), University College Hospital (UCH), Ibadan, Nigeria and its association with socio-demographic, family relationships, spirituality, cognition, depression, functional disability, quality of life, and level of frailty among them.

**Methods:** This will be a cross-sectional descriptive study of 385 older persons (≥60 years) attending the CTAGC, UCH, Ibadan, Nigeria. A semi-structured, interviewer-administered questionnaire will obtain information on the respondents’ demographic, social, economic, family relationships, health profiles, and healthcare utilization patterns. The Sense of Coherence (SOC) status will be measured with Antonovsky’s SOC scale (SOC-13). The information on the respondent’s spirituality, cognition, depression, functional disability, quality of life, family relationship, and level of frailty will be assessed using the spirituality index of well-being, six-item screener, Geriatric depression scale, Barthel’s independence Activities of Daily Living, World Health Organization-Quality of Life brief scale, Sense of Coherence-Family Relations Scale (SCO-FRS), and self-assessment of frailty syndrome, respectively.

**Data analysis:** Data will be entered and analyzed using the Statistical Package for Social Sciences (SPSS) Version 27.0. Tables and charts will be summarised using frequency, proportion, and means. Inferential statistics will test for associations between variables using the Student’s t-test and Analysis of Variance (ANOVA) as appropriate. Linear regression analysis will explore the relationship between significant variables in bivariate analysis with SOC. The level of significance will be set at 5%.

**Implication:** This investigation holds several promises for enhancing psychological well-being, improving physical health outcomes, informing holistic geriatric care, strengthening social support networks, and guiding policy and program development. By prioritizing research and intervention in these areas, we can foster a society that values and supports the well-being of older adults, ensuring they enjoy a fulfilling and dignified life during their golden years.

## Introduction and Rationale

Growing old is a critical stage in life where changes occur both physically, psychologically, and in social relationships. Hence, older adults with health conditions are faced with additional challenges relating to daily lifestyle, reduced social network, and navigating their environments, which potentially either results in the Sense of loss or coherence (Broetje et al., 2019; Ciairano et al., 2008; Jeung et al., 2016). Therefore, as the population of older adults increases across Africa, the importance of enhancing coping resources has been acknowledged in the international literature (Díaz-Castillo & Razo-Gonzalez, 2018; Rohani et al., 2015; Erickson & Lindstrom, 2005). Among older adults, a sense of coherence (SOC) has been proven to be an important influencing factor for coping with varied health challenges (Lovheim et al., 2013; Wang & Kuo, 2006; Helvik et al., 2014). Although evidence suggests that age-long existing traditional family settings contribute to the psychological health of older adults in Nigeria (Akanji et al., 2002; Atchessi et al., 2018; Ajayi et al., 2015; Animasahun & Chapman, 2017; Fajemilehin & Odebiyi, 2011) and the perceived capacity of controlling the reality of ageing, with the desire that life challenges still worth exploring (Adebayo, 2014; Adibe et al., 2022). The combination of these constitutes a high SOC among older adults (Antonovsky, 1979), which needs further exploration within the Nigerian context. In addition, a SOC among older adults is a crucial contributory factor for developing and maintaining mental health and good quality of life (Ciairano et al., 2008; Erikson & Lindstrom, 2005; 2006; Feldt et al., 2003; Geyer, 1997).

Historically, a SOC was conceptualized by Antonovsky (1979) as a global construct that expresses the extent of pervasiveness of an individual capacity to endure through the critical dynamic feelings of confidence that emanate from either predictable internal or external environments. Furthermore, Antonovsky (1987) further describes a SOC in three basic dimensions (manageability, comprehensibility, and meaningfulness). SOC among older adults comes with the perceived Sense that both external and internal environments are considerably predictable and structured. However, validated evidence on the construct of a SOC has not been reported in recent studies among older adults residing in the global south, where the population of older adults with chronic health conditions is increasing exponentially. There is a hope that resources available are designed to fulfil the demands which are required by these stimuli (referred to as manageability) and to consider that these demands warrant spending the energy on (the meaningfulness of engaging in activity) (Lindstrom et al., 2005; Lovheim et al., 2013; Koelen et al., 2017; Wang et al., 2015).

From the life-course perspective, a SOC emerges overtime as age increases, and it is related to good perceived health and quality of life in older adults (Bryant et al., 2001; Holmgren & Soderhamn, 2005; Stenbock-Hult & Sarvimaki, 1994). According to one of Antonovsky’s (1993) lectures in Berkeley, he extensively described the SOC illustrating the transition from adolescence to adulthood and ageing, thus arguing for its usefulness. He positioned an ageing individual into the context of a health continuum from robust health to disease or death and that every individual consistently moves in these directions of the continuum. All these assumptions have been empirically supported in previous research among older adults in the African region. Factors observed to give meaning and purpose to older adults’ status in a longitudinal study on the SOC of older adults in Finland are social relationships, being physically active, being happy with life, and having good health (Takkinen & Ruopila, 2001). Another factor associated with SOC in another study was gender and perceived health status among older adults (Saevareid et al, 2007).

Notably, evidence from the global-north suggests that sustaining a SOC among older adults is strongly dependent on the safety of the community and the ability of the older adults to live in their own homes comfortably, regardless of their income and age (Menec et al., 2011; Center for Disease Control and Prevention, 2020). However, since more than half of older adults in Nigeria reside in rural communities, evidence from Nigeria has not extensively explored the broad importance of building a SOC among older adults residing in rural or urban communities (Daramola et al., 2018; Adebayo, 2014).

One of the critical factors for developing a SOC among older adults is spirituality, which is a complex construct that attempts to search for meaning and provide a reconciliation to an individual’s experiences and personal beliefs (Eriksson, 2022; Peterson, 2000; Polhuis et al., 2020), yet evidence from Nigeria on the spirituality of older adults that intertwine with a sense of coherence has not been reported empirically. Nonetheless, Pate and Bondi (1992) acknowledged that spirituality is the Sense of one’s place in the universe, which gives an individual some context to their ultimate environment regarding their values and belief systems. This implies that, in Africa, the interconnectedness of spirituality and SOC is tied with communication with the deity and other people or a sense that an individual is part of a larger order embedded in the universe.

Despite the availability of studies on the SOC in High-Income Countries (HICs), SOC and its associated factors have been rarely studied in the elderly population of low and middle-income countries, such as Nigeria (Adibe, 2022). A positive but not direct causal relationship has been proposed between the three domains of SOC: comprehensibility, manageability and meaningfulness, and healthy lifestyle. While a direct negative causal could be linked to poor activities of daily living, pain, poor global health status, depression, functional disability, and perceived learned helplessness (Adibe, 2022).

An older person with a high feeling of comprehensibility expects that stimuli/events that appear in the future will be rational, understandable, and predictable, or if they come as surprises, they will be ordered and explicable. Also, an older person with a high feeling of manageability perceives the resources as adequate and available to meet the demands posed by the stimulus and still feels able to cope adequately with the situation. At the same time, an older person with a high feeling of meaningfulness is likelier to feel that life makes sense and that at least some of the problems and demands are worth investing energy in and making commitments for. The two available studies from Nigeria on SOC were on bereavement and grief in adults (Adibe, 2022) and mothers of preschool children (Egbunah et al., 2018). In comparison, none has been carried out among older Nigerians.

## Research Objectives

We aim to determine the Sense of Coherence and associated factors among older adults attending the Chief Tony Anenih Geriatric Centre, University College Hospital, Ibadan, Nigeria.

## Specific objectives

1. To determine the Sense of Coherence (Comprehensibility, Manageability, and Meaningfulness) among older patients presenting at the Geriatric Centre, UCH, Ibadan.
2. To describe the clinical factors such as morbidities, disabilities, functionality, and frailty associated with the Sense of Coherence at the Geriatric Centre, UCH, Ibadan.
3. To explore the influence of family relationships, quality of life, spirituality, religiosity, and beliefs on the Sense of Coherence at the Geriatric Centre, UCH, Ibadan.
4. To determine the predictors of Sense of Coherence among older patients at the Geriatric Centre, UCH, Ibadan.

## Methods and Designs

**Study design:** This will be a cross-sectional study.

**Study setting:** This study will be conducted at the Chief Tony Anenih Geriatric Centre (CTAGC), University College Hospital (UCH), Ibadan. Ibadan is the capital city of Oyo State in the southwestern area of Nigeria and has a population of 3.6 million inhabitants, while Oyo State has 5.6 million people (NPCN, 2006). The CTAGC is a purpose-built centre established on November 17, 2012, to give holistic care to older patients coming to UCH. CTAGC is the pioneer geriatric centre in Nigeria and renders both in-patient and outpatient services.

**Inclusion criteria**: All newly registered male and female patients aged 60 years and above present at the CTAGC Clinic during the study period. The age of the respondents will be determined by asking them, using historical events (Ogunniyi and Osuntokun, 1993; Paraïso et al., 2010), the age at marriage and the age of their first child. Male and female patients aged 60 years and above who presented at the CTAGC, UCH during the study period and met the inclusion criteria will be recruited.

**Exclusion criteria**: All non-consenting and very ill older patients.

### Sample size calculation

The sample size was calculated using Andrew Fisher’s formula for a single mean.

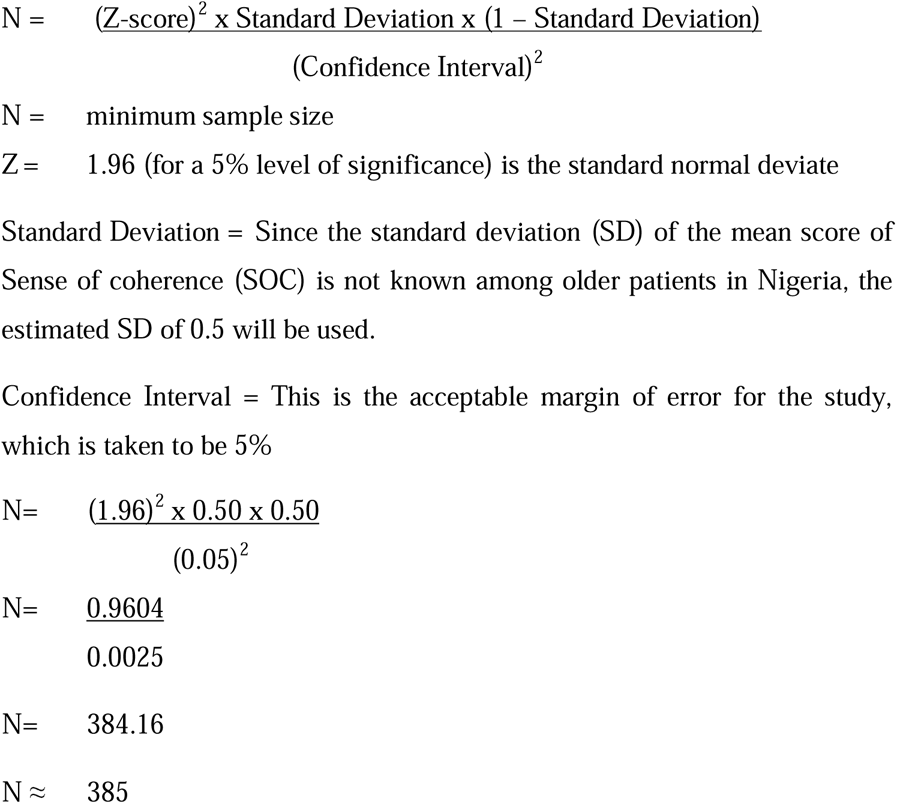

The minimum sample size for this study is three hundred and eighty-five respondents.

**Sampling procedure:** Each month, an average of 230 newly registered older patients are seen monthly at the CTAGC clinic. During the three months of the study, 690 (230 x 3) older patients will be expected to present at the CTAGC Clinic, UCH. Since the sample size is 385, the sample interval is 1.8 (690/385) ≈ 2. Thus, one in every two older patients present at the GOP clinic will be recruited.

**Sampling technique:** This will be by systematic sampling.

### Instruments

This would be an interviewer-administered semi-structured questionnaire.

***Section A:*** will seek information on the respondents’ demographic characteristics such as their age, sex, ethnicity, religion, marital status, and the number of children; socio-economic characteristics like educational level, income, occupation (present and past), living arrangement, lifestyle habits, financial and social support. Information on the previous outpatient visits, previous hospitalization, healthcare utilization pattern, and medication use in the last month will be obtained.

***Section B:*** Validated tools that will be used include the short-form Sense of Coherence (SOC-13) to measure the comprehensibility, manageability, and meaningfulness of the Sense of Coherence and the World Health Organization Quality of Life Brief questionnaire (WHOQoL-Bref) to measure the quality of life. Their spirituality, functional status, cognition, nutritional status, depression, and frailty will be measured with the Spiritual Index of Well-Being (SIWB), Barthel’s Basic Activities of Daily Living (BADL), six-item screener, Mini-Nutritional Assessment short-form (MNA-SF), Sheikh and Yesavage’s Geriatric Depression Scale, and self-reported frailty scale respectively. Additionally, the family relationships will be assessed with the Sense of coherence-family relations.

***Section C:*** The International Classification of Primary Care Diseases (ICPC), which was developed by the World’s Organization of Family Doctors (WONCA) (Adebusoye *et al*., 2011; WONCA, 2011), will be used to seek information on the health problems of the respondents. Anthropometric measurements of height and weight will be done using the standard method, and a complete physical examination will be carried out on the respondents to arrive at the diagnoses.

### Anthropometric measurements

#### Height

Height will be recorded in meters with a measurement stand (stadiometer) manufactured by Seca Corporation, Columbia, Maryland, USA. The stadiometer will be positioned on a flat surface. The patients will be asked to remove their shoes, and their heels will be positioned against the wall. Also, the female patients will be asked to remove the headwear and the hair flattened temporarily with a hard flat surface, making a perpendicular tangent to the wall. The height will be measured to the nearest 0.1 centimetres. For older patients who cannot stand, the knee-length measurement will be used as a proxy for height measurement using Chumlea’s formula (Barcelo et al., 2013; Melo et al., 2014).

#### Weight

Weight will be measured with a weighing scale manufactured by Hana, Japan. Weight will be recorded in kilograms (kg) to the nearest 0.1kg. The weighing scale will be placed on a flat horizontal surface, after which the patients will be asked to remove personal effects like bags, shoes, and heavy clothing. The patient will stand on the scale, and the readings will be made with the researcher standing before the patient. The zero mark will be checked after every reading for accuracy. For those who cannot stand, a weighing chair will be used to measure their weights.

#### Body mass index (BMI)

The BMI of the patients will be calculated from the height in meters and weight in kilograms. BMI is the weight in kilograms divided by height in meters squared, and this will be graded using the WHO anthropometric classification (WHO, 1995; Adebusoye *et al*., 2012). Underweight will be defined as BMI <18.4kg/m^2^ and 18.5 – 24.9 kg/m^2^ as normal. Overweight will be ascribed to elderly patients with a BMI of ≥25.0 kg/m^2^. Overweight will be sub-divided into Pre-obesity BMI 25.0 – 29.9kg/m^2^; Class I obesity as BMI 30.0 to 34.9 kg/m^2^, Class II obesity as BMI 35.0 – 39.9 kg/m^2^, and Class III obesity, which is morbid obesity as BMI of greater than 40.0 kg/m^2^ (WHO, 1995).

#### Blood pressure

The blood pressure will be measured with an Accoson^R^ mercury sphygmomanometer England, calibrated and validated before use. The patients will sit comfortably with their left arm bared and supported at the level of the heart and their feet on the floor. Patients will be allowed to relax, and measurement will start after 5 minutes of rest. Appropriate cuff sizes will be used for each patient, encircling at least 80% of the arm. The appearance of the first sound (Korotkov 1) will be taken as the systolic blood pressure, and the disappearance of the sound (Korotkov 5) as the diastolic blood pressure. The average of two readings separated by 2 minutes will be taken as the blood pressure (WHO/ISH Writing Group, 2003). Hypertension will be staged according to the eighth report of the Joint National Committee on Prevention, Detection, Evaluation, and Treatment of Hypertension (Armstrong, 2014).

### Instruments for Determining the Predictors

#### Sense of Coherence-Short form (SOC-13)

The short form of the SOC will be used in this study (Antonovsky, 1987). The 13-item version includes items measuring each of the three dimensions of SOC. The scale consists of five Comprehensibility items, four Manageability items, and four Meaningfulness items. The original SOC by Antonosky was presented on a 7-point Likert scale (Adibe, 2022). For this study, the short version was modified to a five-scale Likert scale questionnaire with 1=most often (the worst possible option) to 5=never (the best possible option) (Egbunah, Uti, and Sofola, 2018) will be used. All items will then be summed to create a total SOC score ranging from 13 to 65. A higher SOC score indicates a greater SOC (Egbunah, Uti, and Sofola, 2018). Five items are negatively stated and reversed in scoring, so a high score always indicates a stronger SOC (Egbunah, Uti, and Sofola, 2018). The short-form SOC (SOC-13) has alpha reliabilities between 0.74 and 0.91 (Adibe, 2022). The Soc-13 has been used in Nigeria with a reported Cronbach alpha of 0.89 for the overall scale and subscale Cronbach alpha of 0.77 for Comprehensibility, 0.87 for Manageability, and 0.64 for Meaningfulness (Adibe, 2022).

#### The Family Relationships

The family relationship will be assessed using the Sense of Coherence-Family Relations scale (Chiang and Lee, 2018). This is a three-item scale that measures family involvement, emotional ties, and family sociability:

i. “How much are your significant family members involved in your daily life?” (Involvement)
ii. “How are your emotional relationships with your close family members?” (Emotional ties)
iii. “What is the atmosphere of your family interactions?” (sociability).

The responses are made on 5 points Likert scale: (i) “none” or “bad”; (ii) “sometimes” or “fair”; (iii) “often” or “good”; (iv) “usually” or “great”; and (v) “always” or “perfect”. The possible scores ranged from 5-15, with a higher score indicating more positive family relations. The Cronbach’s alpha was reported as 0.73, the composite reliability was 0.74, and the average variance extracted was 50% (Chiang and Lee, 2018).

#### World Health Organisation Quality of Life Brief Questionnaire (WHOQoL-Bref)

The World Health Organization Quality of Life instrument (WHOQoL-Bref) will be used to measure the quality of life of the subjects. The WHOQOL-Bref is a cross-culturally applicable tool developed by the WHOQOL Group in 1998 for the subjective evaluation of health-related QoL (Gureje *et al*., 2008). It is a valid measure of QoL in older people (Naumann and Byrne, 2004; Gureje *et al*., 2008). The WHOQOL-Bref is designed as a self-rating instrument that can also be interviewer-administered with excellent internal reliability (Cronbach alpha = 0.86) and has four domains: physical, psychological, social, and environmental (Naumann and Byrne, 2004; Gureje et al., 2008).

#### Spiritual Index of Well-Being (SIWB)

The spiritual quality of life of the respondents will be measured with the spiritual index of well-being (SIWB). The SIWB has two domains– 6 items on the intrapersonal self-efficacy domain and six on the life scheme domain. It is measured on a 5-point Likert scale ranging from 1 (strongly disagree) to 5 (strongly agree). Higher scores indicated lower levels of spiritual well-being (Frey, 2004; Wu, Yang, and Koo, 2017).

#### Barthel’s Basic Activities of Daily Living (BADL)

The functional disability of the respondents will be assessed using Barthel’s activities of daily living index. The Barthel Index is a 10-item simple-to-administer tool for assessing self-care and mobility activities of daily living. It is widely used in geriatric assessment settings (Mahoney and Barthel, 1965). The reliability, validity, and overall utility of the Barthel Index are rated as good to excellent (Mahoney and Barthel, 1965). Information for assessing functional disability among older people is gained from observation, self-report, or informant report. Total possible scores range from 0 – 20, with lower scores indicating increased functional disability (Mahoney and Barthel, 1965; Ajayi et al., 2015).

#### The Six-item screener

Cognition will be screened with ‘the six-item screener’ (Callahan et al., 2002). This is a brief and reliable instrument for identifying subjects with cognitive impairment, and its diagnostic properties are comparable to the full MMSE (sensitivity 95.2 and specificity 86.7) (Callahan et al., 2002). It is easily scored by a simple summation of errors. The sensitivity and specificity of the six-item screener for a diagnosis of dementia were 88.7 and 88.0, respectively (Callahan et al., 2002).

#### Mini-nutritional assessment-short-form (MNA-SF)

Mini-nutritional assessment-short-form (MNA-SF), used in Nigerian studies, will screen for malnutrition among the subjects (Nestle Nutrition Institute, 2009; Nzeagwu and Uwaegbute, 2010; Adebusoye et al., 2012).

#### Geriatric Depression Scale (GDS)

The Geriatric Depression Scale (GDS) developed by Sheikh and Yesavage will be used to assess depression (Sheikh and Yesavage, 1988). The GDS-short form, which had been used in Nigerian studies (Akosile *et al*., 2018; Awunor et al., 2018) and the same scoring system will be used.

#### Self-reported frailty scale

The self-reported frailty scale will be applied to the respondents or their proxy. It consists of dichotomous questions directly related to each component of the frailty phenotype, which is considered the gold standard model: unintentional weight loss, fatigue, low physical activity, decreased physical strength, and decreased walking speed. The score will be classified as not frail (no component identified), pre-frail (presence of one or two components), and frail (presence of three or more components). The sensitivity and specificity for identifying pre-frail individuals were reported as 89.7% and 24.3%, respectively, while those for identifying frail individuals were 63.2% and 71.6%, respectively (Nunes et al., 2015).

#### Three-item Perception of illness scale

The respondents’ self–reported health would be assessed and scored using the ‘three-item perception of illness scale’. Each item is graded from 1 to 5 (poor to excellent), which are aggregated into a composite score of perception of health (score = 3 to 15) (Idler and Kasp, 1995). The perception of illness scale has a Cronbach’s alpha coefficient of 0.65 (Exavery, Klipstein-grobusch, and Debpuur, 2013).

##### Translation of Instruments

The researchers will administer the questionnaire in English and interpret it into the Yoruba language (the local dialect of most respondents) when necessary. The questionnaire will take about 35 minutes to be administered, and measurements will take about 30 minutes.

#### Consent for the Study

Approval for the study will be obtained from the Director of the CTAGC, UCH, Ibadan, and the University of Ibadan/University College Hospital Institutional Ethical Review Board (UI/UCH IRB). Informed consent from each respondent will be obtained before the examination and administration of the questionnaire.

## Data analysis

At the end of each study day, the administered questionnaires will be sorted out, cross-checked after each interview, and coded serially. Data will be entered and analyzed using the Statistical Package for Social Sciences (SPSS) Version 27.0. Tables and charts will be summarised using frequency, proportion, and means. Appropriate charts will be used to illustrate categorical variables. Inferential statistics will test for associations between variables using the Student’s t-test and Analysis of Variance (ANOVA) as appropriate. Linear regression analysis will explore the relationship between significant variables in bivariate analysis with SOC. The level of significance will be set at 5%.

### Ethical Considerations

Ethical clearance for the study will be obtained from the joint University of Ibadan/University College Hospital Ethical Review Board. Permission will also be sought from the Director, Chief Tony Anenih Geriatric Centre (CTAGC), UCH. Ibadan. Written informed consent will be obtained from each participant before data collection. The respondent will either sign or thumb-print depending on their level of literacy.

### Confidentiality of Data

The names of participants will not be on the questionnaire to maintain confidentiality. Only serial numbers allocated to the participants will be written on the questionnaire. The names, hospital numbers, and identification codes will be recorded in a separate notebook accessible only to the researcher and the assistant to easily retrieve the results of physical findings and investigations as the respondents may request. They will be assured that their responses will be kept confidential. The questionnaires will be kept safely in a locked cupboard. Data entered on the computer will be pass-worded and accessible only to the researcher, data entry clerk, and statistician.

### Translation of protocol to the prominent local language for easy communication

The questionnaire will be translated into Yoruba, the local language, back-translated to English, and field-tested to ensure the original meaning is retained.

### Beneficence to the participants

All respondents will be managed for their primary complaints. The study’s outcome will be useful in focused counselling and appropriate intervention for the respondents.

### Non-maleficence to the participants

This study is not harmful to the participant, as confidentiality will be ensured. The consulting rooms set apart for the interview will ensure the privacy of the respondents.

### Right to decline/withdraw from the study without loss of benefits (Voluntariness)

The participants are free to decide not to participate and could choose to discontinue at any point during the interview process without jeopardizing their opportunity to be treated or attended to.

## Supporting information

Questionnaire

## Data Availability

All data produced in the present study will be made available upon reasonable request to the authors.

## Acknowledgments

The second author is a recipient of the Social Science and Humanities Research Council (SSHRC) 2022 Vanier Canada Doctoral Scholarship

## Notes

### Competing Interest Statement

The authors have declared no competing interest.

### Funding Statement

This study did not receive any funding

