## Supplementary material for "Determinants of Sense of Coherence among older adults attending a Geriatric Centre in Nigeria A study Protocol": Questionnaire

#### SECTION A: PERSONAL INFORMATION

**DATE __________________________________________**

**SERIAL NUMBER: ______________________**

**1.	SEX**	Male 1	Female 2

**2.	AGE as at last birthday**______________________________________

**3.	MARITAL STATUS**

(a) Married	(b) Single	(c) Divorced	(d) Widowed	(e) Separated

**4.	RELIGION**

(a) Christianity	(b) Islam	(c) Traditional	(d) others

**5.	ETHNIC GROUP**

(a) Yoruba	(b) Ibo	(c) Hausa	(d) Others, please specify____________

**6.	EDUCATIONAL LEVEL**

(a) None	(b) Primary school	(c) Secondary school	(d) Tertiary

**7.	OCCUPATION**

At present_________________________________

If retired, what was your occupation? ____________________________

**8.	YOUR INCOME** (Naira per month) ___________________

**9.	With whom are you living?**

(a) Alone (b) With spouse	(c) With children /Grandchildren (d) With relatives and friends

**10.	Number of Children (Alive)**_____________________________

**11.	Who supports you financially?**

(a) Self	(b) Spouse	(c) Children / Grandchildren

(d) Other relatives	(e) Friends

**12.	Who supports you socially?**

(a) Self	(b) Spouse	(c) Children / Grandchildren

(d) Other relatives	(e) Friends

**13.	When were you first admitted into a hospital as an in-patient?**

(a) Never been admitted	(b) Before the age of 60 years

(c) On reaching or after the age of 60years	(d) Don’t know

**14.	How many times have you been admitted to the hospital as an in-patient, on or after reaching the age of 60 years?**...............................................................

**15.	How many times have you visited the hospital as an outpatient on or after reaching the age of 60 years?..............................................................................**

##### 16. Where did you receive treatment for your ailments in the last year?

##### **(a) clinics (b) chemists (c) Traditional settings (d) Self-medication**

##### 17. SOCIAL HABITS

| **Do you take it?** | **YES** | **NO** | **QUIT** |
| --- | --- | --- | --- |
| (a) Alcohol | If yes, how many bottles per week?  For how long? |  |  |
| (b) Tobacco | If yes, how many sticks per day?  For how long? |  |  |
| (c) Cannabis | If yes, how many wraps per day?  For how long? |  |  |
| (d) Coffee | If yes, how many cups per day?  For how long? |  |  |

##### 18. Please rate the level of your physical activity

(i) Not active	(ii) Moderately active	(iii) Very active

**19. Do you have longevity in your family?**	**Yes - 1	No – 2**

**SECTION B:**

**20.	Sense of Coherence- Short form (SOC-13)**

***Instructions-*** Please indicate how much you agree or disagree with each by entering a number alongside it according to the following code:

**1= Most often	2=Always;	3=Sometimes;	4=Rarely;	5=Never**

|  |  | **1** | **2** | **3** | **4** | **5** |
| --- | --- | --- | --- | --- | --- | --- |
|  | **Comprehensibility** |  |  |  |  |  |
| 1 | Has it happened in the past that you were surprised by the behaviour of people whom you thought you knew well (R)? |  |  |  |  |  |
| 2 | Do you have the feeling that you are in an unfamiliar situation and don’t know what to do? |  |  |  |  |  |
| 3 | Do you have the feeling that you are in an unfamiliar situation and don’t know what to do? |  |  |  |  |  |
| 4 | Do you have very mixed-up feelings and ideas? |  |  |  |  |  |
| 5 | When something happened, have you generally found that? |  |  |  |  |  |
|  | **Manageability** |  |  |  |  |  |
| 6 | Has it happened that people whom you counted on disappointed you (R)? |  |  |  |  |  |
| 7 | Do you have the feeling that you’re being treated unfairly? |  |  |  |  |  |
| 8 | Many people – even those with strong character – sometimes feel like sad sacks (losers) in certain situations. How often have you felt this way in the past (R)? |  |  |  |  |  |
| 9 | How often do you have feelings that you’re not sure you can keep things under control? |  |  |  |  |  |
|  | **Meaningfulness** |  |  |  |  |  |
| 10 | Do you have the feeling that you don’t really care about what goes on around you? |  |  |  |  |  |
| 11 | Until now your life has had no clear goals or purpose at all |  |  |  |  |  |
| 12 | Doing the things, you do every day is a source of pain and boredom (R). |  |  |  |  |  |
| 13 | How often do you have the feeling that there’s little meaning in the things you do in your daily life? |  |  |  |  |  |

**21. SOC-Family Relationship Index**

**1= “none” or “bad”	2= “sometimes” or “fair”	3= “often” or “good”**

**4= “usually” or “great”	5= “always” or “perfect”**

|  |  | **1** | **2** | **3** | **4** | **5** |
| --- | --- | --- | --- | --- | --- | --- |
| 1 | How much are your significant family members involved in your daily life?” (Involvement) |  |  |  |  |  |
| 2 | “How are your emotional relationships with your close family members?” (Emotional ties) |  |  |  |  |  |
| 3 | “What is the atmosphere of your family interactions?” (sociability). |  |  |  |  |  |

**22. WHOQOL-BREF**

The following questions ask how you feel about your quality of life. I will read out each question to you, along with the response options. Please choose the answer that appears most appropriate. If you are unsure about which response to give to a question, the first response you think of is often the best one (The numbers after responses indicate the scores of the responses).

Please keep in mind your standards, hopes, pleasures, and concerns. We ask that you think about your life in the last four weeks (The overall quality of life and general health facet).

|  |  | Very poor | Poor | Neither poor nor good | Good | Very good |
| --- | --- | --- | --- | --- | --- | --- |
| 1. | How would you rate your quality of life? | 1 | 2 | 3 | 4 | 5 |

|  |  | Very dissatisfied | Dissatisfied | Neither satisfied nor dissatisfied | Satisfied | Very satisfied |
| --- | --- | --- | --- | --- | --- | --- |
| 2. | How satisfied are you with your life? | 1 | 2 | 3 | 4 | 5 |

The following questions ask about how much you have experienced certain things in the last four weeks.

|  |  | Not at all | A little | A moderate amount | Very much | An extreme amount |
| --- | --- | --- | --- | --- | --- | --- |
| 3. | To what extent do you feel that physical pain prevents you from doing what you need to do? | 5 | 4 | 3 | 2 | 1 |
| 4. | How much do you need any medical treatment to function in your daily life? | 5 | 4 | 3 | 2 | 1 |
| 5. | How much do you enjoy life? | 1 | 2 | 3 | 4 | 5 |
| 6. | To what extent do you feel your life to be meaningful? | 1 | 2 | 3 | 4 | 5 |

|  |  | Not at all | A little | A moderate amount | Very much | Extremely |
| --- | --- | --- | --- | --- | --- | --- |
| 7. | How well are you able to concentrate? | 1 | 2 | 3 | 4 | 5 |
| 8. | How safe do you feel in your daily life? | 1 | 2 | 3 | 4 | 5 |
| 9. | How healthy is your physical environment? | 1 | 2 | 3 | 4 | 5 |

The following questions ask about how completely you experienced or were able to do certain things in the last four weeks.

|  |  | Not at all | A little | Moderately | Mostly | completely |
| --- | --- | --- | --- | --- | --- | --- |
| 10 | Do you have enough energy for everyday life? | 1 | 2 | 3 | 4 | 5 |
| 11 | Are you able to accept your bodily appearance? | 1 | 2 | 3 | 4 | 5 |
| 12 | Have you enough money to meet your needs? | 1 | 2 | 3 | 4 | 5 |
| 13 | How available to you is the information that you need in your day-to-day life? | 1 | 2 | 3 | 4 | 5 |
| 14 | To what extent do you have the opportunity for leisure activities? | 1 | 2 | 3 | 4 | 5 |

|  |  | Very poor | Poor | Neither poor nor good | Good | Very good |
| --- | --- | --- | --- | --- | --- | --- |
| 15 | How well are you able to get around? | 1 | 2 | 3 | 4 | 5 |

|  |  | Very dissatisfied | Dissatisfied | Neither satisfied nor dissatisfied | Satisfied | Very satisfied |
| --- | --- | --- | --- | --- | --- | --- |
| 16 | How satisfied are you with your sleep? | 1 | 2 | 3 | 4 | 5 |
| 17 | How satisfied are you with your ability to perform your daily living activities? | 1 | 2 | 3 | 4 | 5 |
| 18 | How satisfied are you with your capacity for work? | 1 | 2 | 3 | 4 | 5 |
| 19 | How satisfied are you with yourself? | 1 | 2 | 3 | 4 | 5 |
| 20 | How satisfied are you with your relationships? | 1 | 2 | 3 | 4 | 5 |
| 21 | How satisfied are you with your sex life? | 1 | 2 | 3 | 4 | 5 |
| 22 | How satisfied are you with the support you get from your friends? | 1 | 2 | 3 | 4 | 5 |
| 23 | How satisfied are you with the conditions of your living place? | 1 | 2 | 3 | 4 | 5 |
| 24 | How satisfied are you with your access to health services? | 1 | 2 | 3 | 4 | 5 |
| 25 | How satisfied are you with your transport? | 1 | 2 | 3 | 4 | 5 |

The following question refers to how often you have felt or experienced certain things in the last four weeks.

|  |  | **Never** | **Seldom** | **Quite often** | **Very often** | **Always** |
| --- | --- | --- | --- | --- | --- | --- |
| **26** | How often do you have negative feelings such as blue mood, despair, anxiety, and depression? |  |  |  |  |  |

### 24. The Spirituality Index of Well-Being

|  | Statement | Strongly agree | Agree | Neither Agree nor Disagree | Disagree | Strongly Disagree |
| --- | --- | --- | --- | --- | --- | --- |
| 1 | There is not much I can do to help myself. |  |  |  |  |  |
| 2 | Often, there is no way I can complete what I have started. |  |  |  |  |  |
| 3 | I can’t begin to understand my problems. |  |  |  |  |  |
| 4 | am overwhelmed when I have personal difficulties and  problems. |  |  |  |  |  |
| 5 | I don’t know how to begin to solve my problems. |  |  |  |  |  |
| 6 | There is not much I can do to make a difference in my life. |  |  |  |  |  |
| 7 | I haven’t found my life’s purpose yet. |  |  |  |  |  |
| 8 | don’t know who I am, where I came from, or where I am going |  |  |  |  |  |
| 9 | I have a lack of purpose in my life |  |  |  |  |  |
| 10 | In this world, I don’t know where I fit in. |  |  |  |  |  |
| 11 | I am far from understanding the meaning of life. |  |  |  |  |  |
| 12 | There is a great void in my life at this time. |  |  |  |  |  |

### 25. Barthel’s Independence in Activities of Daily Living

|  | **With help** | **Independent** |
| --- | --- | --- |
| 1. Feeding (if food needs to be cut up = help) |  |  |
| 2. Moving from wheelchair to bed and return (includes sitting up in bed) |  |  |
| 3. Personal toilet (wash face, comb hair, shave, clean teeth) |  |  |
| 4. Getting on and off the toilet (handling clothes, wiping, flushing) |  |  |
| 5. Bathing self |  |  |
| 6. Walking on a level surface (or if unable to walk, propel wheelchair) |  |  |
| 7. Ascend and descend stairs |  |  |
| 8. Dressing (includes tying shoes and fastening fasteners) |  |  |
| 9. Controlling bowels |  |  |
| 10. Controlling bladder |  |  |

**26.	SCREENER FOR COGNITION (6-ITEM)**

|  | **Incorrect (0)** | **Correct (1)** |
| --- | --- | --- |
| 1. What year is this? |  |  |
| 2. What month is this? |  |  |
| 3. What is the day of the week |  |  |
| **Tell the respondent to repeat items (4 to 6) after you. Then, ask the respondents to repeat the items after 10 minutes** | | |
| 4. Orange |  |  |
| 5. Table |  |  |
| 6. Naira |  |  |

**27.	MNA-SF (Malnutrition screening)**

**A. Has food intake declined over the past three months due to loss of appetite, digestive problems, chewing or swallowing difficulties?**

**0** = severe decrease in food intake	**1** = moderate decrease in food intake

**2** = no decrease in food intake

**B. Weight loss during the last three months**

**0** = weight loss greater than 3 kg (6.6 lbs)	**1** = does not know

**2** = weight loss between 1 and 3 kg (2.2 and 6.6 lbs)	**3** = no weight loss

**C. Mobility**

**0** = bed or chair bound	**1** = able to get out of bed/chair but does not go out

**2** = goes out

**D. Has suffered psychological stress or acute disease in the past three months?**

**0** = yes	**2** = no

**E. Neuropsychological problems**

**0** = severe dementia or depression	**1** = mild dementia

**2** = No psychological problems

**F. Body Mass Index (BMI)**

**0** = BMI less than 19	**1** = BMI 19 to less than 21

**2** = BMI 21 to less than 23	**3** = BMI 23 or greater

**28.	GERIATRIC DEPRESSION SCALE**

Choose the best answer for how you have felt over the past week:

|  | **Items** | **YES** | **NO** |
| --- | --- | --- | --- |
| 1 | Are you basically satisfied with your life? |  |  |
| 2 | Do you often get bored? |  |  |
| 3 | Do you often feel helpless? |  |  |
| 4 | Do you prefer to stay at home rather than going out and doing new things? |  |  |
| 5 | Do you feel pretty worthless the way you are now? |  |  |

**29. Self-reported assessment of frailty syndrome**

|  |  | **Yes** | **No** |
| --- | --- | --- | --- |
| 1 | In the last 12 months, did you lose weight without going on any diet? |  |  |
| 2 | In the last 12 months, do you feel weaker or think your strength has decreased? |  |  |
| 3 | Do you think that you are walking more slowly than you did 12 months ago? |  |  |
| 4 | Do you think that you are currently performing less physical activity than you did 12 months ago? |  |  |
| 5 | In the past week, did you feel that you could not perform daily activities (you started something but could not finish)? |  |  |

**30.	Self-rated health**

|  |  | **Excellent** | **Good** | **Fair** | **Poor** | **Bad** |
| --- | --- | --- | --- | --- | --- | --- |
| 1 | How would you rate your health at the present time? |  |  |  |  |  |
| 2 | How would you rate your health 6 months ago? |  |  |  |  |  |
| 3 | How would you rate your health compared with your age-mate? |  |  |  |  |  |

**31.	ANTHROPOMETRIC MEASUREMENTS**

Height (Meter)________________________________

Weight (kg)___________________________________

**32. Diagnoses**

a) **_______________________________**
b) **_______________________________**
c) **_______________________________**
d) **_______________________________**
